# Vaginal dinoprostone versus placebo for pain relief during intrauterine device insertion: a systematic review and meta-analysis of randomized controlled trials

**DOI:** 10.1101/2020.10.08.20209239

**Authors:** Ahmed Abu-Zaid, Majed S. Alshahrani, Nisreen A. Albezrah, Najlaa T. Miski, Saud A. Aboudi, Mohammed Abuzaid, Osama Alomar, Hany Salem, Ismail A. Al-Badawi, Saeed Baradwan

## Abstract

**Objective:** To investigate the safety and efficacy of vaginal dinoprostone versus placebo in pain relief during intrauterine device (IUD) insertion.

**Design:** Systematic review and meta-analysis of randomized placebo-controlled trials.

**Setting:** Not applicable.

**Patient(s):** Women undergoing IUD insertion and receiving vaginal dinoprostone or placebo.

**Intervention(s):** PubMed, Scopus, Web of Science, and Cochrane Library were screened from inception to 01-October-2020, using the following search strategy: (dinoprostone OR cervidil OR prepidil) AND (intrauterine device OR iud).

**Main outcome measure(s):** IUD insertion related pain, patient satisfaction, provider ease of IUD insertion, and side effects.

**Result(s):** Five studies met the study inclusion criteria, comprising 862 patients; equally 431 patients received vaginal dinoprostone and placebo. All studies had an overall low risk of bias. When compared to placebo, dinoprostone significantly correlated with decreased pain at tenaculum placement (SMD=−0.79, 95% CI [−1.43, −0.16], p=0.01), decreased pain at uterine sounding (SMD=−0.88, 95% CI [−1.54, −0.22], p=0.009), decreased pain at IUD insertion (SMD=−1.18, 95% CI [−1.74, −0.61], p<0.001), decreased need for additional analgesia (RR=0.34, 95% CI [0.22, 0.53], p<0.001), increased patient satisfaction (SMD=1.41, 95% CI [0.62, 2.20], p<0.001), and increased provider ease of IUD insertion (SMD=−1.17, 95% CI [−1.62, −0.73], p<0.001). Fever was statistically significantly higher in dinoprostone versus placebo group (RR=3.73, 95% CI [1.47, 9.44], p=0.006). All other side effects—including nausea, vomiting, shivering, diarrhea, abdominal cramps, vasovagal attack, uterine perforation, and postprocedural bleeding—did not substantially differ between both groups.

**Conclusions:** This first ever meta-analysis advocates that dinoprostone is safe, effective, and yields favorable analgesic outcomes during IUD insertion.

## Introduction

Intrauterine devices (IUDs) deliver a largely effective, harmless, and long-lasting method of reversible contraception with an analogous efficacy to tubal sterilization (1, 2). The use of IUDs is highly endorsed by the American College of Obstetricians and Gynecologists (3) and American Academy of Pediatrics (4) to avoid unplanned pregnancies among sexually active adolescents and young women. Globally, the two most frequently utilized IUDs comprise the levonorgestrel-releasing intrauterine system (LNG-IUS) and copper-containing intrauterine device (Cu-IUD) (1, 2), both of which are equally used in nulliparous and multiparous women (5).

There are a few downsides associated with IUD use. Importantly, pain perception is one of the substantial factors contributing to restricted utility of IUDs, particularly among nulliparous women who possess relatively narrower uterine cavities when compared to their multiparous counterparts (6, 7). This remark is in line with the perspective that a large proportion of healthcare personnel restrict IUD administration to nulliparous women owing to worries pertaining to anticipated insertion pain and procedural difficulties (8-10). Indeed, each step of IUD insertion procedure can instigate a large deal of pain perception (11). Therefore, proper control of pain before, during, and after IUD insertion is critically important to favorably maximize the IUD usage frequency and minimize the rate of unplanned pregnancies among sexually active adolescents and young women.

The optimal method of pain relief during IUD insertion remains undefined (12, 13). A contemporary systematic review and network meta-analysis of numerous lines of pharmacologic analgesic interventions—including placebo, nonsteroidal anti-inflammatory drugs, nitric oxide donors, lavender scent, lidocaine, and misoprostol—demonstrated no tangible effectiveness for IUD insertion-related pain (14). Conversely, lidocaine-prilocaine cream (genital mucosal application) was the most effective intervention for IUD insertion-related pain (14). However, when lidocaine-prilocaine was compared head-to-head with other pharmacologic interventions, including placebo, it did not exhibit significantly reduced pain 5-20 minutes after IUD insertion (14). More research is warranted for alternative analgesics to control pain during IUD insertion procedure.

Dinoprostone is a naturally occurring prostaglandin E2 equivalent (15). It is frequently employed in obstetrics for labor induction, which is mediated through cervical ripening and prompting of uterine contractions with an equivalent labor-inducing efficacy to misoprostol but substantially less adverse events (16). Additionally, dinoprostone has been exploited successfully prior to diagnostic hysteroscopy to ease the procedure and reduce associated pain without considerable toxicity when compared to placebo or misoprostol (17, 18). Only a very limited number of trials examined the safety and efficacy of vaginal dinoprostone versus placebo in facilitating IUD insertion and decreasing its related pain (12, 18-21). To date, no meta-analysis has been conducted to amass the data and inform concrete conclusions. Therefore, the aim of this study is to systematically and meta-analytically synthesize evidence from randomized controlled trials that scrutinized the safety and efficacy of vaginal dinoprostone versus placebo for pain relief among women undergoing IUD insertion.

## Methods

This systematic review and meta-analysis was conducted in harmony with the Preferred Reporting Items for Systematic Reviews and Meta-Analyses (PRISMA) guidelines (22).

### Literature search strategy

Four databases (PubMed, Scopus, Web of Science, and Cochrane Library) were screened from inception to October 1st, 2020. The following search strategy was used for all databases: (dinoprostone OR cervidil OR prepidil) AND (intrauterine device OR iud). There was no language restriction.

### Inclusion and exclusion criteria

We included all articles that met the following criteria for our PICOS evidence-based research question: (I) Patients: women who received LNG-IUS or Cu-IUD for contraception, (II) Intervention: vaginal dinoprostone, (III) Comparator: vaginal placebo, (IV) Outcomes: efficacy and safety endpoints, and (V) Study design: randomized controlled trials. We excluded drugs other than dinoprostone, indications other than contraception, non-randomized study designs, non-human trials, abstracts, and articles without full texts.

### Screening of Results

The retrieved citations were exported using EndNote software and duplicates were crossed out. The screening of results was completed through a two-fold phase. The first phase involved title and abstract screening of all citations. The second phase involved retrieval of the full text of all potential citations. Moreover, the reference lists of the included studies were reviewed manually for potential inclusion of other relevant studies. Two authors screened the citations independently and disagreements were resolved by a consensus among the two authors.

### Risk of bias assessment of the included studies

We utilized the Cochrane’s risk of bias tool in evaluating the quality of the included studies (23). This tool is elaborated in the Cochrane Handbook for Systematic Reviews of Interventions, Version 5.1.0, Chapter 8. This tool appraises the following domains: random sequence generation, allocation concealment, blinding of participants and personnel, blinding of outcome assessment, incomplete outcome data, selective outcome reporting, and other potential sources of bias. Two authors evaluated the risk of bias independently and disagreements were resolved by a consensus among the two authors.

### Data Extraction

We extracted three types of data: (I) baseline characteristics of the included studies, (II) efficacy outcomes, and (III) safety outcomes. Data about baseline characteristics of the included studies comprised author’s first name, year of publication, national clinical trial identifier, country, type of IUD device, study group, sample size, timing of drug administration before IUD procedure, and the person administering the drug. Moreover, patients’ age, body mass index, parity, gravidity, position of uterus, history of previous abortion, and history of previous IUD were extracted. Data about efficacy outcomes included pain at tenaculum placement, pain at uterine sounding, pain at IUD insertion, pain after IUD insertion (10-30 mins), need for additional analgesia after procedure, duration of procedure, ease of IUD insertion as reported by healthcare providers, and procedural satisfaction as reported by patients. Pain scores were evaluated according to the 10-cm/100-mm visual analogue scale (VAS) in which “0” corresponded to no pain at all and “10 cm/100 mm” corresponded to the worst possible pain imaginable. Likewise, ease of IUD insertion as reported by healthcare providers was scored according to a 10-cm/100-mm VAS-like metric in which “0” corresponded to easy insertion and “10 cm/100 mm” corresponded to extremely difficult insertion. Equally, procedural satisfaction as reported by patients was scored according to a 10-cm/100-mm VAS-like metric in which “0” corresponded to no satisfaction and “10 cm/100 mm” corresponded to maximum satisfaction. Data about safety outcomes included nausea, vomiting, diarrhea, shivering, fever, abdominal cramps, postprocedural bleeding, vasovagal attack, and uterine perforation. Four authors participated in data extraction and verification.

### Data analysis

Review Manager Software version 5.4 was used for meta-analysis. Continuous data were analyzed using the inverse variance method and reported as weighted mean difference (WMD) or standardized mean difference (SMD), as appropriate, with 95% confidence interval (95% CI). Dichotomous data were analyzed using the Mantel–Haenszel method and reported as risk ratio (RR) with 95% CI. Statistical heterogeneity was established if chi-square was p<0.1 and I-square test (I^2^) was >50% (24). Fixed- and random-effects models were used for meta-analysis of homogenous and heterogeneous data, respectively. Publication bias was not evaluated since the number of included studies (n=5) was lower than the minimum required (n=10) (27).

## Results

### Search results and summary of included studies

Literature search generated a total of 67 studies after omission of duplicated ones. After title and abstract screening, 50 studies were excluded and the remaining 17 studies progressed to full text screening for eligibility. Finally, a total of five (n=5) studies met the inclusion criteria and were included in the qualitative and quantitative synthesis (12, 13, 19-21). **Supplemental Figure 1** displays the PRIMSA flowchart. This meta-analysis included 862 patients; equally 431 patients received vaginal dinoprostone and placebo. All studies originated in Egypt. Three studies included nulliparous women, whereas one study included only patients who delivered by cesarean section. Furthermore, three studies included patients receiving Cu-IUD as the method of contraception. The dose of vaginal dinoprostone was consistent (3 mg) in all studies, however, the duration of drug application differed between studies, ranging from 2 to 12 hours before procedure. Drugs were administered by nurses in three studies and self-administered in two studies. The baseline characteristics of the included studies are displayed in **Supplemental Table 1**.

### Risk of bias assessment of the included studies

Overall, the included studies showed an overall low risk of bias. In two studies, the drugs (vaginal dinoprostone and placebo) were self-administered by patients. Although measures had been taken by investigators to remind patients about the time to vaginally self-administer the drugs three (19) and 12 (13) hours before the procedure, however, this cannot be certainly established, and we judged the other bias domain as high risk. The graph and summary of risk of bias are depicted in **Supplemental Figure 2**.

### Efficacy outcome: Pain at tenaculum insertion

Three studies were meta-analyzed (12, 13, 19). The overall effect estimate revealed significantly reduced pain at tenaculum insertion in the dinoprostone versus placebo group (SMD=−0.79, 95% CI [−1.43, −0.16], p=0.01). Pooled analysis was heterogeneous (I^2^=88%, p=0.0002) (**Figure 1A**).

**Figure 1.**
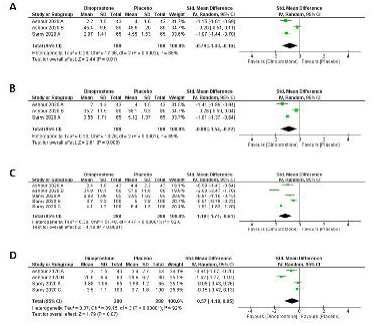
Forest plot showing the effect size for (A) pain at tenaculum insertion, (B) pain at uterine sounding, (C) pain at intrauterine device (IUD) insertion, and (D) pain after IUD insertion (10-30 mins) between vaginal dinoprostone and placebo groups.

### Efficacy outcome: Pain at uterine sounding

Three studies were meta-analyzed (12, 13, 19). The overall effect estimate revealed significantly reduced pain at uterine sounding in the dinoprostone versus placebo group (SMD=−0.88, 95% CI [−1.54, −0.22], p=0.009). Pooled analysis was heterogeneous (I^2^=89%, p<0.001) (**Figure 1B**).

### Efficacy outcome: Pain at IUD insertion

Five studies were meta-analyzed (12, 13, 19-21). The overall effect estimate revealed significantly reduced pain at IUD insertion in the dinoprostone versus placebo group (SMD=−1.18, 95% CI [−1.74, −0.61], p<0.001). Pooled analysis was heterogeneous (I^2^=92%, p<0.001) (**Figure 1C**).

### Efficacy outcome: Pain after IUD insertion (10-30 mins)

Four studies were meta-analyzed (12, 13, 19, 21). The overall effect estimate did not exhibit statistically significant difference between both groups with regard to pain after IUD insertion (SMD=−0.57 [−1.19, 0.05], p=0.07). Pooled analysis was heterogeneous (I^2^=92%, p<0.001) (**Figure 1D**).

### Efficacy outcome: Need for additional analgesia

Four studies were meta-analyzed (12, 13, 19, 21). The overall effect estimate revealed significantly reduced need for additional analgesia in the dinoprostone versus placebo group (RR= 0.34, 95% CI [0.22, 0.53], p<0.001). Pooled analysis was homogenous (I^2^=0%, p=0.88) (**Figure 2**).

**Figure 2.**
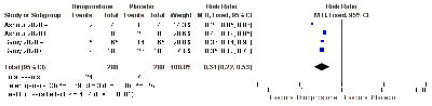
Forest plot showing the effect size for need for additional analgesia after intrauterine device (IUD) insertion between vaginal dinoprostone and placebo groups.

### Efficacy outcome: Duration of IUD insertion procedure

Five studies were meta-analyzed (12, 13, 19-21). The overall effect estimate did not exhibit statistically significant difference between both groups (MD= −0.49 [−1.15, 0.18], p=0.15). Pooled analysis was heterogeneous (I^2^=96%, p<0.001) (**Supplemental Figure 3**).

### Efficacy outcome: Patient satisfaction

Four studies were meta-analyzed (12, 13, 19, 21). The overall effect estimate revealed significantly increased patient satisfaction in the dinoprostone versus placebo group (SMD=1.41, 95% CI [0.62, 2.20], p<0.001). Pooled analysis was heterogeneous (I^2^=94%, p<0.001) (**Figure 3**).

**Figure 3.**
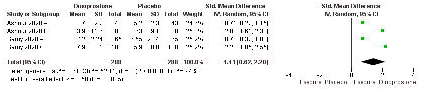
Forest plot showing the effect size for patient satisfaction between vaginal dinoprostone and placebo groups.

### Efficacy outcome: Ease of IUD insertion according to the healthcare provider

Five studies were meta-analyzed (12, 13, 19-21). The overall effect estimate revealed significantly increased ease of IUD insertion in the dinoprostone versus placebo group (SMD=−1.17, 95% CI [−1.62, −0.73], p<0.001). Pooled analysis was heterogeneous (I^2^=88%, p<0.001) (**Figure 4**).

**Figure 4.**
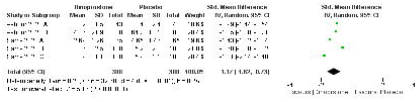
Forest plot showing provider ease of insertion of intrauterine device (IUD) between vaginal dinoprostone and placebo groups.

### Safety outcomes

Five studies were meta-analyzed (12, 13, 19-21). Frequency of fever was statistically significantly higher in the dinoprostone versus placebo group (RR=3.73, 95% CI [1.47, 9.44], p=0.006). Pooled analysis was homogenous (I^2^=0%, p=0.92). All other side effects—including nausea (RR=1.03, 95% CI [0.69, 1.53], p=0.90), vomiting (RR=2.11, 95% CI [0.97, 4.61], p=0.06), diarrhea (RR=2.78, 95% CI [0.95, 8.09], p=0.06), shivering (RR=2.38, 95% CI [0.96, 5.90], p=0.06), abdominal cramps (RR=1.76, 95% CI [0.73, 4.26], p=0.21), and postprocedural bleeding (RR=1.02, 95% CI [0.92, 1.14], p=0.72)—did not substantially differ between both groups. Pooled analyses were homogenous for all side effects (I^2^=0%), except abdominal cramps; pooled analysis was heterogeneous (I^2^=85%, p<0.001). No cases of vasovagal attack or uterine perforation happened in either group (**Supplemental File 1**).

## Discussion

This systematic review and meta-analysis endeavored to examine the safety and efficacy of vaginal dinoprostone versus placebo in controlling pain during IUD insertion. We included five high quality randomized controlled trials comprising 862 patients (dinoprostone, n=431 and placebo, n=431).

Despite IUD insertion is relatively a quick (5-10 minutes) procedure, each step of it can bring about variable extents of pain (11). Sources of pain comprise speculum insertion, tenaculum placement, transcervical sounding of uterus, forward advancement of IUD inserter into uterine cavity, and postprocedural pain. When compared to placebo, our pooled analyses showed that dinoprostone significantly reduced pain at tenaculum placement, transcervical sounding of uterus, and IUD insertion. This clinically meaningful pain reduction was positively correlated with increased patient-reported procedural satisfaction and decreased requirement for additional postprocedural analgesia. Nonetheless, although pain after IUD insertion was reduced, this pain reduction did not reach statistical significance. The lack of statistically significant pain reduction after IUD insertion could be credited to the variable time points that were used to assess this parameter across all the pooled four studies (10, 15, 20, and 30 min).

Our findings indicated that dinoprostone was correlated with significantly increased ease of IUD insertion by healthcare providers. Nonetheless, the IUD insertion time was not substantially impacted. This increased ease of insertion score could be ascribed to the favorable cervical ripening effects of dinoprostone (15, 28). Mechanistically, dinoprostone initiates cervical softening through stimulation of interleukin 8 (IL-8), which in turn facilitates influx of polymorphonuclear leukocyte neutrophils that orchestrate remodeling of cervical extracellular matrix and induction of progesterone withdrawal (28).

There are some risk factors that may subscribe to more severe pain perception during IUD insertion. Such factors comprise nulliparity, women who delivered only by cesarean section, and use of LNG-IUS (25, 26, 29). When compared to parous women, nulliparous women relatively possess narrower uterine cavities which may not properly fit the dimensions of conventional IUDs, thus culminating in higher IUD insertion pain (7). Moreover, while a history of cesarean section does not automatically preclude IUD insertion, nevertheless, a structurally disfigured uterus secondary to repetitive cesarean section may potentially correlate with difficult IUD insertion and discomfort (30). Lastly, the comparatively thicker diameter of LNG-IUS inserter (ranging from 4.65 to 4.85 mm) contrasted to Cu-IUD counterpart (around 4.0 mm) may cause more pain during insertion (29). In our study, subgroup analyses according to parity and device of contraception (data not shown) revealed that dinoprostone equally significantly correlated with better pain control, patient satisfaction, and ease of IUD insertion during the procedure when compared to placebo. This study suggests that dinoprostone can yield successful insertion of IUD in both nulliparous and parous women with favorable efficacy outcomes.

Our summary data depicted that side effects did not significantly differ between dinoprostone and placebo groups. Fever was the only drug-related side effect that was significantly higher in the dinoprostone group when compared to placebo group. This side effect is, to a larger degree, expected as the association between prostaglandin E2 and occurrence of fever is well documented in literature (31). Other than fever which can be adequately and conservatively managed by antipyretics, our study suggests dinoprostone is highly effective and associated with satisfactory safety profile.

Dinoprostone is widely used for the obstetric indication of labor induction owing to favorable cervical softening and uterine contractility properties (15, 28). A meta-analysis demonstrated that while dinoprostone is efficaciously equivalent to misoprostol in labor induction, it is associated with better toxicity profile, particularly in terms of lower frequencies of tachysystole and hypertonic uterine dysfunction (16). Also, use of dinoprostone has been extended to non-obstetric indications such as hysteroscopy in nulliparous (17) and postmenopausal (18) women and correlated with better pain control and ease of procedure when compared to placebo or misoprostol. This study further supports the clinical utility of dinoprostone in an additional non-obstetric indication—that is, during IUD insertion to control procedure-related pain and facilitate its ease of conduction.

The optimal method of pain relief during IUD insertion remains undefined (12, 13). A contemporary systematic review and network meta-analysis of numerous lines of pharmacologic analgesic interventions showed that lidocaine-prilocaine cream (genital mucosal application) was the most effective intervention for controlling IUD insertion-related pain (14). Dinoprostone was not included in the aforementioned network meta-analysis. Thus, future research should be geared toward direct comparison of efficacy and safety of vaginal dinoprostone versus lidocaine-prilocaine cream (and other active comparators such as misoprostol, lidocaine, and nonsteroidal anti-inflammatory drugs) in controlling pain associated with IUD insertion. This should be achieved through development of well-designed randomized controlled trials that take into account women who are at high risk for more painful experiences, such as nulliparous women, women who delivered only by cesarean section, women who failed previous insertions, and women who will receive the comparatively thicker LNG-IUS (as opposed to Cu-IUD) devices.

Dinoprostone possesses two major drawbacks that ought to be acknowledged. First, from a financial perspective, it is costly when compared to its closely related comparator misoprostol (32, 33). Second, from a physiochemical perspective, it is unstable at room temperature and should be stored in freezer/refrigerator until before use to preserve its potency (12).

This study has several strengths. First, this is the first systematic review and meta-analysis that pooled the efficacy and safety of vaginal dinoprostone versus placebo in controlling pain during IUD insertion. We included only randomized placebo-controlled trials (n=5) to lessen potentials of bias and confounding in our pooled conclusions. All included studies were of high quality and low risk of bias. We strictly adhered to PRISMA guidelines during the conduction of this research. Moreover, we reported many efficacy and safety endpoints. All in all, our study suggests the beneficial role of vaginal dinoprostone for a non-obstetric indication, which is controlling pain during IUD insertion. Nonetheless, this study has some limitations that should be recognized. Such limitations comprise the subjective evaluation of pain which may be impacted by the patients’ sociodemographics or pre-anxiety levels. The studies varied substantially with regard to timing of vaginal dinoprostone administration, ranging from two to 12 hours prior to IUD insertion, as well as timing of assessing pain after IUD insertion, ranging from 10 to 30 mins. All these variations could have negatively impacted the factual assessment of the efficacy endpoint. Importantly, the optimal timing of dinoprostone administration is yet to be determined. Lastly, all the included studies originated from one country/institute, thus relatively limiting the generalizability of the outcomes.

## Conclusions

This systematic review and meta-analysis suggests that vaginal dinoprostone is correlated with increased ease of insertion by providers, higher satisfaction by patients, and decreased IUD insertion-related pain. Moreover, dinoprostone is largely safe with very tolerable toxicity profile. All in all, this study supports the clinical utility of vaginal dinoprostone for pain relief during IUD insertion, including those nulliparas who are at high risk for increased pain perception.

## Data Availability

All relevant data are included in the article and/or its supplementary information files.

## Acknowledgement

None

## Supplemental Tables

Supplemental Table 1. Baseline characteristics of the included studies.

## Supplemental Figures

Supplemental Figure 1. Preferred Reporting Items for Systematic Reviews and Meta-Analyses (PRISMA) flowchart.

Supplemental Figure 2. Risk of bias summary and graph.

Supplemental Figure 3. Forest plot showing the effect size for duration of intrauterine device (IUD) insertion between vaginal dinoprostone and placebo groups.

## Supplemental Files

Supplemental File 1. Forest plots showing the effect sizes for side effects between vaginal dinoprostone and placebo groups.

