## Supplementary figures and images for "Vaginal dinoprostone versus placebo for pain relief during intrauterine device insertion: a systematic review and meta-analysis of randomized controlled trials"

### Supplemental Figure 1

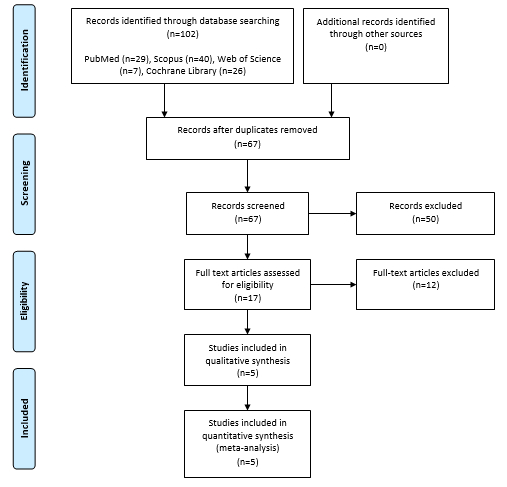

### Supplemental Figure 2

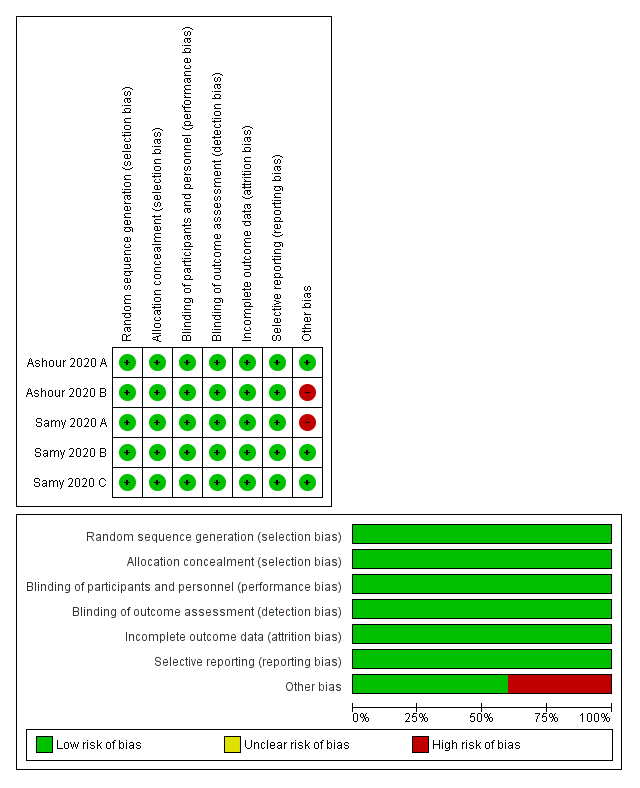

### Supplemental Figure 3

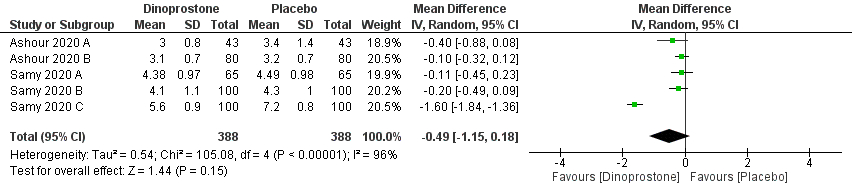
