## Supplemental File 1 for "Vaginal dinoprostone versus placebo for pain relief during intrauterine device insertion: a systematic review and meta-analysis of randomized controlled trials"

**Figure 1**. Forest plot showing the overall effect estimate between vaginal dinoprostone and placebo groups regarding the side effect of fever.

| | Dinoprostone | | Placebo | | | Risk Ratio |
|---|---|---|---|---|---|---|
| **Study or Subgroup** | **Events** | **Total** | **Events** | **Total** | **Weight** | **M-H, Fixed, 95% CI** |
| Ashour 2020 A | 2 | 43 | 1 | 43 | 18.2% | 2.00 [0.19, 21.24] |
| Ashour 2020 B | 3 | 80 | 1 | 80 | 18.2% | 3.00 [0.32, 28.23] |
| Samy 2020 A | 6 | 65 | 2 | 65 | 36.4% | 3.00 [0.63, 14.32] |
| Samy 2020 B | 5 | 100 | 0 | 100 | 9.1% | 11.00 [0.62, 196.33] |
| Samy 2020 C | 4 | 100 | 1 | 100 | 18.2% | 4.00 [0.46, 35.16] |
| **Total (95% CI)** | | **388** | | **388** | **100.0%** | **3.73 [1.47, 9.44]** |
| Total events | 20 | | 5 | | | |

Heterogeneity: Chi² = 0.92, df = 4 (P = 0.92); I² = 0%
Test for overall effect: Z = 2.78 (P = 0.006)

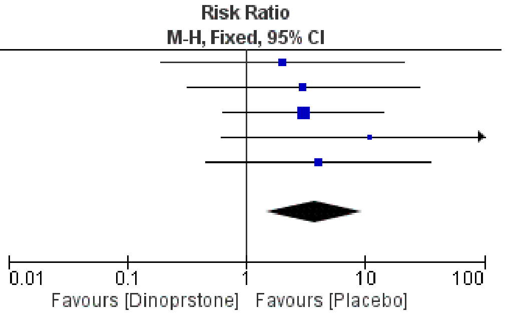


**Figure 2**. Forest plot showing the overall effect estimate between vaginal dinoprostone and placebo groups regarding the side effect of nausea.

| | Dinoprostone | | Placebo | | | Risk Ratio |
|---|---|---|---|---|---|---|
| **Study or Subgroup** | **Events** | **Total** | **Events** | **Total** | **Weight** | **M-H, Fixed, 95% CI** |
| Ashour 2020 A | 3 | 43 | 2 | 43 | 5.1% | 1.50 [0.26, 8.53] |
| Ashour 2020 B | 4 | 80 | 2 | 80 | 5.1% | 2.00 [0.38, 10.61] |
| Samy 2020 A | 4 | 65 | 3 | 65 | 7.7% | 1.33 [0.31, 5.72] |
| Samy 2020 B | 21 | 100 | 30 | 100 | 76.9% | 0.70 [0.43, 1.14] |
| Samy 2020 C | 8 | 100 | 2 | 100 | 5.1% | 4.00 [0.87, 18.37] |
| **Total (95% CI)** | | **388** | | **388** | **100.0%** | **1.03 [0.69, 1.53]** |
| Total events | 40 | | 39 | | | |

Heterogeneity: Chi² = 6.38, df = 4 (P = 0.17); I² = 37%
Test for overall effect: Z = 0.12 (P = 0.90)

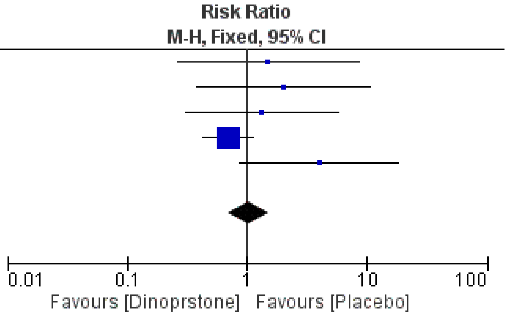


**Figure 3**. Forest plot showing the overall effect estimate between vaginal dinoprostone and placebo groups regarding the side effect of vomiting.

| | Dinoprostone | | Placebo | | | Risk Ratio |
|---|---|---|---|---|---|---|
| **Study or Subgroup** | **Events** | **Total** | **Events** | **Total** | **Weight** | **M-H, Fixed, 95% CI** |
| Ashour 2020 A | 2 | 43 | 1 | 43 | 11.1% | 2.00 [0.19, 21.24] |
| Ashour 2020 B | 4 | 80 | 2 | 80 | 22.2% | 2.00 [0.38, 10.61] |
| Samy 2020 A | 3 | 65 | 2 | 65 | 22.2% | 1.50 [0.26, 8.68] |
| Samy 2020 B | 5 | 100 | 3 | 100 | 33.3% | 1.67 [0.41, 6.79] |
| Samy 2020 C | 5 | 100 | 1 | 100 | 11.1% | 5.00 [0.59, 42.04] |
| **Total (95% CI)** | | **388** | | **388** | **100.0%** | **2.11 [0.97, 4.61]** |
| Total events | 19 | | 9 | | | |

Heterogeneity: Chi² = 0.89, df = 4 (P = 0.93); I² = 0%
Test for overall effect: Z = 1.88 (P = 0.06)

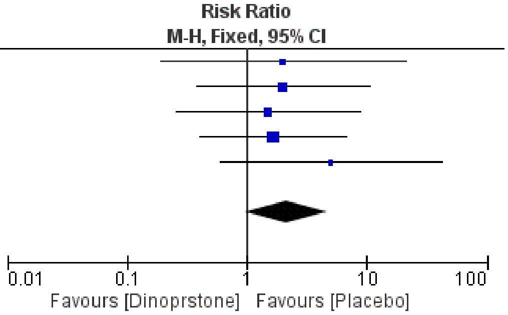

**Figure 4**. Forest plot showing the overall effect estimate between vaginal dinoprostone and placebo groups regarding the side effect of diarrhea.

| Study or Subgroup | Dinoprostone Events | Dinoprostone Total | Placebo Events | Placebo Total | Weight | Risk Ratio M-H, Fixed, 95% CI |
|---|---|---|---|---|---|---|
| Ashour 2020 A | 1 | 43 | 1 | 43 | 22.2% | 1.00 [0.06, 15.48] |
| Ashour 2020 B | 2 | 80 | 1 | 80 | 22.2% | 2.00 [0.19, 21.62] |
| Samy 2020 A | 2 | 65 | 1 | 65 | 22.2% | 2.00 [0.19, 21.52] |
| Samy 2020 B | 4 | 100 | 0 | 100 | 11.1% | 9.00 [0.49, 165.00] |
| Samy 2020 C | 3 | 100 | 1 | 100 | 22.2% | 3.00 [0.32, 28.35] |
| **Total (95% CI)** | | **388** | | **388** | **100.0%** | **2.78 [0.95, 8.09]** |
| Total events | 12 | | 4 | | | |

Heterogeneity: Chi² = 1.31, df = 4 (P = 0.86); I² = 0%
Test for overall effect: Z = 1.87 (P = 0.06)

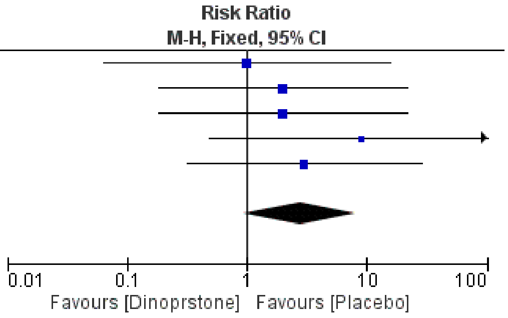


**Figure 5**. Forest plot showing the overall effect estimate between vaginal dinoprostone and placebo groups regarding the side effect of shivering.

| Study or Subgroup | Dinoprostone Events | Dinoprostone Total | Placebo Events | Placebo Total | Weight | Risk Ratio M-H, Fixed, 95% CI |
|---|---|---|---|---|---|---|
| Ashour 2020 A | 1 | 43 | 1 | 43 | 15.4% | 1.00 [0.06, 15.48] |
| Ashour 2020 B | 3 | 80 | 2 | 80 | 30.8% | 1.50 [0.26, 8.74] |
| Samy 2020 A | 3 | 65 | 2 | 65 | 30.8% | 1.50 [0.26, 8.68] |
| Samy 2020 B | 5 | 100 | 0 | 100 | 7.7% | 11.00 [0.62, 196.33] |
| Samy 2020 C | 3 | 100 | 1 | 100 | 15.4% | 3.00 [0.32, 28.35] |
| **Total (95% CI)** | | **388** | | **388** | **100.0%** | **2.38 [0.96, 5.90]** |
| Total events | 15 | | 6 | | | |

Heterogeneity: Chi² = 2.04, df = 4 (P = 0.73); I² = 0%
Test for overall effect: Z = 1.88 (P = 0.06)

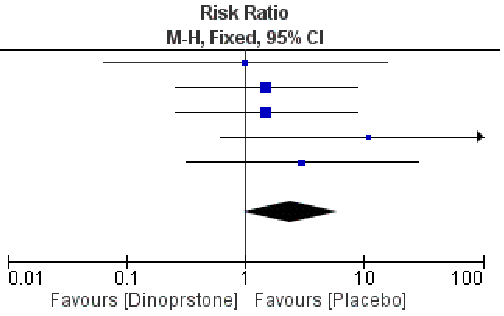


**Figure 6**. Forest plot showing the overall effect estimate between vaginal dinoprostone and placebo groups regarding the side effect of abdominal cramps.

| Study or Subgroup | Dinoprostone Events | Dinoprostone Total | Placebo Events | Placebo Total | Weight | Risk Ratio M-H, Random, 95% CI |
|---|---|---|---|---|---|---|
| Ashour 2020 A | 6 | 43 | 3 | 43 | 16.3% | 2.00 [0.53, 7.49] |
| Ashour 2020 B | 12 | 80 | 5 | 80 | 19.3% | 2.40 [0.89, 6.50] |
| Samy 2020 A | 9 | 65 | 4 | 65 | 18.0% | 2.25 [0.73, 6.94] |
| Samy 2020 B | 95 | 100 | 93 | 100 | 25.5% | 1.02 [0.95, 1.10] |
| Samy 2020 C | 15 | 100 | 8 | 100 | 21.0% | 1.88 [0.83, 4.22] |
| **Total (95% CI)** | | **388** | | **388** | **100.0%** | **1.76 [0.73, 4.26]** |
| Total events | 137 | | 113 | | | |

Heterogeneity: Tau² = 0.80; Chi² = 27.57, df = 4 (P < 0.0001); I² = 85%
Test for overall effect: Z = 1.25 (P = 0.21)

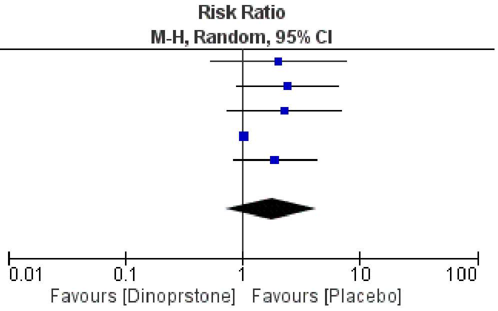

**Figure 7**. Forest plot showing the overall effect estimate between vaginal dinoprostone and placebo groups regarding the side effect of postprocedural bleeding.

| | Dinoprostone | | Placebo | | | Risk Ratio |
|---|---|---|---|---|---|---|
| **Study or Subgroup** | **Events** | **Total** | **Events** | **Total** | **Weight** | **M-H, Fixed, 95% CI** |
| Ashour 2020 B | 2 | 80 | 4 | 80 | 4.0% | 0.50 [0.09, 2.65] |
| Samy 2020 B | 93 | 100 | 91 | 100 | 91.9% | 1.02 [0.94, 1.11] |
| Samy 2020 C | 6 | 100 | 4 | 100 | 4.0% | 1.50 [0.44, 5.15] |
| **Total (95% CI)** | | **280** | | **280** | **100.0%** | **1.02 [0.92, 1.14]** |
| Total events | 101 | | 99 | | | |

Heterogeneity: Chi² = 1.08, df = 2 (P = 0.58); I² = 0%
Test for overall effect: Z = 0.36 (P = 0.72)

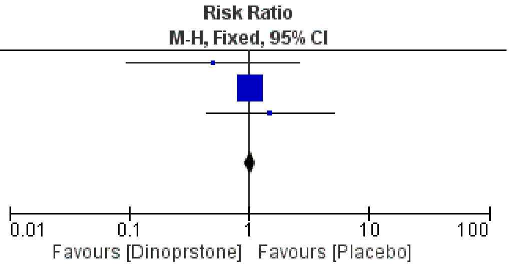
