## Supplemental Table 1 for "Vaginal dinoprostone versus placebo for pain relief during intrauterine device insertion: a systematic review and meta-analysis of randomized controlled trials"

Table 1. Baseline characteristics of the included studies.

| Study identifier | Ref | Country | IUD device | Population | Group | n | Parity  Mean ± SD | Gravidity  Mean ± SD | Age (years)  Mean ± SD | BMI (kg)  Mean ± SD | History of previous abortion  n (%) | Normal position of uterus  n (%) | Previous IUD insertion  n (%) | Timing of drug administration before procedure (hours), person |
| --- | --- | --- | --- | --- | --- | --- | --- | --- | --- | --- | --- | --- | --- | --- |
| Ashour 2020 A  NCT04080336 | (12) | Egypt | Cu-IUD | Nulliparous | Dinoprostone | 43 | NR | 0.2 ± 0.8 | 20.9 ± 2.2 | 28.2 ± 5.2 | 4 (9.3) | 33 (76.7) | NR | 3, self |
|  |  |  |  |  | placebo | 43 | NR | 0.4 ± 1.0 | 20.4 ± 1.7 | 29.7 ± 6.3 | 8 (18.6) | 32 (74.4) | NR |  |
| Ashour 2020 B  NCT04046302 | (19) | Egypt | Cu-IUD | Parous | Dinoprostone | 80 | 2.7 ± 1.0 | NR | 28.6 ± 4.6 | 25.0 ± 3.0 | 35 (43.8) | 65 (81.3) | 16 (20) | 3, nurse |
|  |  |  |  |  | placebo | 80 | 2.5 ± 1.1 | NR | 29.4 ± 4.5 | 24.3 ± 3.0 | 30 (37.5) | 59 (73.8) | 13 (16.3) |  |
| Samy 2020 A  NCT04079140 | (13) | Egypt | LNG-IUS | Nulliparous | Dinoprostone | 65 | NR | 0.62 ± 0.78 | 20.51 ± 1.13  20.62 ± 1.11 | 28.26 ± 2.97  29.26 ± 3.22 | 28 (43.1) | 31 (47.7) | NR  NR | 12, nurse |
|  |  |  |  |  | placebo | 65 | NR | 0.68 ± 0.77 |  |  | 32 (49.2) | 35 (53.8) |  |  |
| Samy 2020 B  NCT04079140 | (20) | Egypt | Cu-IUD | Nulliparous | Dinoprostone | 100 | NR | NR | 26.6 ± 6.3 | 21.9 ± 2.2 | 21 (21) | 82 (82) | NR | 6, self |
|  |  |  |  |  | placebo | 100 | NR | NR | 26.9 ± 6.9 | 22.1 ± 3.9 | 17 (17) | 78 (78) | NR |  |
| Samy 2020 C  NCT04045548 | (21) | Egypt | LNG-IUD | Cesarean delivery only | Dinoprostone | 100 | 2.4 ± 1.0 | NR | 28.8 ± 4.5 | 25.3 ± 4.2 | NR | 73 (73) | 19 (19) | 2, nurse |
|  |  |  |  |  | placebo | 100 | 2.6 ± 1.1 | NR | 28.0 ± 5.5 | 26.3 ± 4.1 | NR | 77 (77) | 17 (17) |  |

BMI: body mass index; Cu-IUD: copper-containing intrauterine device; LNG-IUS: levonorgestrel-releasing intrauterine system, NCT: national clinical trial identifier (clinicaltrials.gov); NR: not reported; Ref: reference; SD: standard deviation
